# A Classification Approach for Predicting COVID-19 Patient’s Survival Outcome with Machine Learning Techniques

**DOI:** 10.1101/2020.08.02.20129767

**Authors:** Abdulhameed Ado Osi, Mannir Abdu, Usman Muhammad, Auwalu Ibrahim, Lawan Adamu Isma’il, Ahmad Abubakar Suleiman, Hasssan Sarki Abdulkadir, Safiya Sani Sada, Hussaini Garba Dikko, Muftahu Zubairu Ringim

## Abstract

COVID-19 is an infectious disease discovered after the outbreak began in Wuhan, China, in December 2019. COVID-19 is still becoming an increasing global threat to public health. The virus has been escalated to many countries across the globe. This paper analyzed and compared the performance of three different supervised machine learning techniques; Linear Discriminant Analysis (LDA), Random Forest (RF), and Support Vector Machine (SVM) on COVID-19 dataset. The best level of accuracy between these three algorithms was determined by comparison of some metrics for assessing predictive performance such as accuracy, sensitivity, specificity, F-score, Kappa index, and ROC. From the analysis results, RF was found to be the best algorithm with 100% prediction accuracy in comparison with LDA and SVM with 95.2% and 90.9% respectively. Our analysis shows that out of these three classification models RF predicts COVID-19 patient’s survival outcome with the highest accuracy. Chi-square test reveals that all the seven features except sex were significantly correlated with the COVID-19 patient’s outcome (P-value < 0.005). Therefore, RF was recommended for COVID-19 patient’s outcome prediction that will help in early identification of possible sensitive cases for quick provision of quality health care, support and supervision.

## 1. Introduction

In December, 2019 an outbreak of pneumonia of unknown cause emerged in Wuhan, Hubei Province, China that led to the discovery of coronavirus. Since then, the disease has been spreading rapidly to more than 150 countries across the globe with more than 3,132,000 confirmed cases of infection and about 230,000 deaths as at 30^th^ April,2020 [1]. In February 2020 the World health organization termed this newly discovered disease as COVID-19 (coronavirus 2019) and subsequently declared it as pandemic on 12^th^ March, 2020 [2]. COVID-19 also known as Severe Acute Respiratory Syndrome Coronavirus 2 (SARS-CoV-2) is a communicable disease that cause respiratory infection that is generally similar to Severe Acute Respiratory Syndrome (SARS-CoV) and Middle East Respiratory Syndrome (MERS-CoV) in severe cases. The disease can be transmitted from person to person via a droplets coming out from the mouth or nose of infected person. The most common symptoms of COVID-19 are: fever; fatigue and dry cough [3]. Some patients may have sore throat, diarrhea, headache and nasal congestion. However, most COVID-19 patients don’t develop any symptoms (asymptotic).

Current studies have shown that most of the COVID-19 patients with underline comorbidity and elderly patients are more likely to develop severe cases which may require ICU care and could lead to death [4]. Several existing literature reported diabetes and hypertension as the most common comorbidities for people with COVID-19 [3] [5] [4] [6]. Although, clinical results showed that being diabetic or hypertensive has no association with the risk of contracting the diseases [7]. However, it could increase chances for severe and critical COVID-19 conditions [8] [3].

Currently, the ongoing pandemic (COVID-19) has attracted the interest of many researchers. Different statistical models have been used in many studies to predict COVID-19 cases, prevalence and mortality [9]used Multivariate Logistic Regression to determine the risk factor associated with mortality, [3] used Cox Regression Models to explore potential risk factors associated with clinical outcomes, While [10] applied Chi-square Test and Spearman’s Ranked Correlation to explored the significant differences between severe and non-severe COVID-19 cases using the clinical and laboratory characteristic of hospitalized patients. [11] used the Mann Whitney U test or Kruskal-Wallis test for comparison significance differences for univariate and chi-square for categorical variables, and then modeled patient’s survival with Hierarchical Cox Regression to identify poor outcome risk population. [12] applied Chi-square to test the association between being HFABP positive and COVID-19 severity using epidemiological, clinical, and laboratory characteristics.

Machine Learning is modern data analysis technique deals with patterns and correlations to extract useful information from the raw data. Machine Learning played a significant role in fighting the previous epidemic, more especially in the area of the disease diagnosis, medication, prediction and drug/vaccine development [13]. Numerous Machine Learning Algorithms have shown powerful prediction capabilities in the diseases prediction [14] [15] [16] [17]. Machine learning technique has recently gained attention for the COVID-19 outbreak spreading prediction [18].. There is gap in the literature for studies dedicated to COVID-19, most of the existing studies are limited to cases prediction. This study focuses on the application of supervised machine learning techniques (i.e SM, RF, LDA) to predict the survival outcome of COVID-19 patients based on some demographic, clinical and epidemiological characteristics. Also to assess the performance of the methods using seven different metrics. The paper equally investigates the effect of these demographics and clinical variables to the patient’s outcome.

## 2. Material and Methods

### 2.1 Dataset and variables

This study uses detail information of patients’ demographic, epidemiological, clinical characteristics (including age, sex, comorbidities, hospital length of stay, travel history, signs, and symptoms), and their definite survival outcomes (discharge or death) available at https://drive.google.com/file/d/1bYcMAd-lOiSlOVGl5qxG1Sq1auyzmWHZ/view. A total sample of 425 confirmed of COVID-19 patients were extracted and who were died or discharged on or before 11^th^ March, 2020 were included in the study. This study was approved by the General Research Ethics Committee of the Kano University of Science and Technology, Wudil, Nigeria. The data were partitioned into two parts (training and test data set), the first part (80%) was used to develop the learning algorithms while the second part (20%) to test the performances of the algorithms.

### 2.2 Prediction models

Three different types of classification models: Linear Discriminant Analysis (LDA), Random Forest (RF), and Support Vector Machines (SVM) were used. These methods have been widely reported and demonstrated as successful methods for classification [17]. Description of each of these classification models is given below.

#### 2.2.1 Linear Discriminant Analysis

Discriminant Analysis (DA) find a set of prediction equation based on independent variables that are used to classify individuals. The main objectives of DA are to find a predictive equation for classifying new individuals and interpreting the predictive equation for a better understanding of the relationship among the variables.

Linear Discriminant Analysis (LDA) formulated by [19] is most commonly used DA. The original Linear Discriminant Analysis was described for a 2-class problem,, and it was later generalized to multiple class by [20]. LDA makes some assumptions about the dataset: the data is Gaussian, and each attribute has the same variance.

LDA use of Bayes theorem to construct the decision rule. Let *π_k_* represent the prior probability of k^th^ class, and let *f_k_*(*X*) *≡* Pr(*X* = *x|Y = k*) denote a conditional density of X given that the observation comes from k^th^ class. The posterior probability is given as

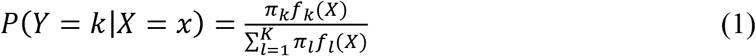

By the two LDA assumptions

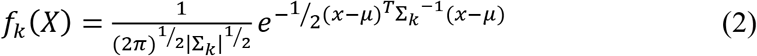

Where ∑*_k_* = ∑∀*_k_*

The decision rule is to assign an observation X=x to the class for which

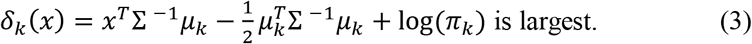

#### 2.2.2 Support Vector Machine

The Support Vector Machine (SVM) is a powerful technique for general (nonlinear supervised machine learning algorithm that is suitable for linear and non-linear problems and can be used for both regression and classification. The aim of SVM is to find a hyperplane in a multidimensional space that splits the feature space into two distinct groups. A new subject is then classified based on which side of the hyperplane he lies on. SVM uses kernel function to converts non-separable problems into separable problems by mapping the training data into higher-dimensional feature space, then finds the hyperplane that maximizes the distance from the nearest subjects and achieves maximum separation. The kernel function can be linear, polynomial or radial, and the choice of the kernel can have a large effect on model outputs. SVM method has successfully achieved high predictive performance in health care researches. In this study, we implemented SVM using the “e1071” package in the R statistical environment.

#### 2.2.3 Random forest

Random Forests (RF) developed by [21] are among the most widely used machine learning algorithm. In RF a “forest” of decision trees are generated on different bootstrap samples by re-sampling the training data that limit. for each single decision tree a random subset of features is randomly chosen at each node to decide optimal split. For classification problems, the ensemble of simple trees votes for the most popular class. The response of each tree depends on a set of predictor values chosen independently (with replacement) and with the same distribution for all trees in the forest, which is a subset of the predictor values of the original data set. The Random Forest has two important parameters, the number of predictive variables to randomly choose at each node for splitting (mtry) and the number of trees to grow in the forest (ntree) [21]. In this study we used the R package “randomForest”, with 1000 trees and mtry = 3, which is square root of the total number of predictor variables.

### 2.3 Performance evaluation for the classification methods

Model performance evaluation is a core part of constructing effective machine learning model. The evaluation can be performed with a set of performance measure metrics, most of whom are derived from confusion matrix. The confusion matrix compares the original (observed) class with predicted class. It consists of four possible outcomes; TP stands for true positives TN (true negatives), FP (false positives), and FN (false negatives). TP and TN are representing a situation whereby the classifier correctly classified positive and negative (“Died” and “Discharged” in our case) instances respectively. FP denotes number of cases where the model misclassified positive as negative while FN represents patients negative incorrectly classified as positive. Table 1 shows layout of the confusion matrix.

**Table1:**
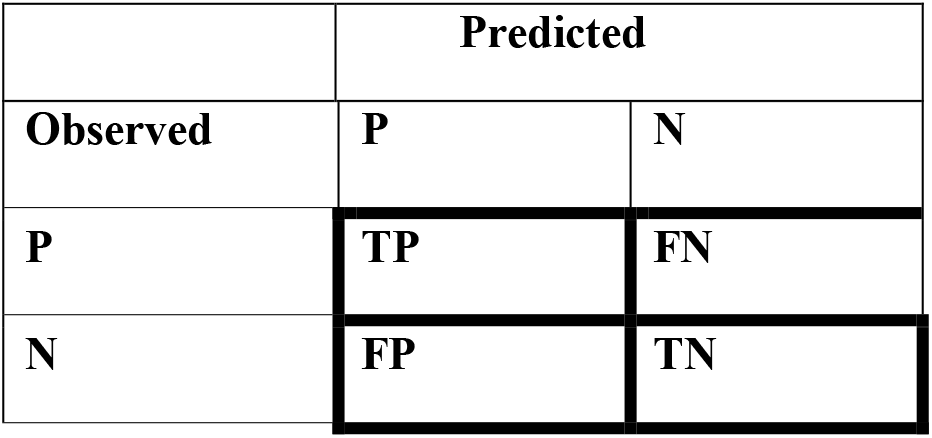
Confusion matrix Predicted.

| Observed | P | N |
| --- | --- | --- |
| P | TP | FN |
| N | FP | TN |

Generally, there is no universally accepted metric for measuring classifiers performance. However, the choice of metric depends on the application. Care need to be taken on how choose the evaluation metrics as it is obvious that, at the same time a classifier could perform very well on a particular metric and badly on the another. Therefore, correct selection of performance metrics is essential to achieve reliable results. [22] suggested that, a classifier should be evaluated using set of performance metrics that ate not closely related to reduce redundancy. In this study, we employed seven most commonly evaluation metrics of two different types; threshold metric (precision, accuracy, sensitivity, specificity, F-score and kappa) and rank metric(AUC-ROC). The metrics are defined below.

i. ***Accuracy:*** Accuracy is a widely used metric for monitoring machine learning models performance and defines as the ration of the correctly classified instances to total number of instances, for unbalanced data this metric can be misleading.

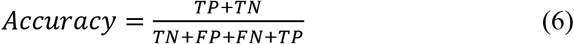
ii. ***F- score:*** The F-score is a single-value metric based on two parameters. It is weighted average of the precision and recall values. In other word, is the harmonic mean of the precision and recall. The range of F-score is between 0 and 1 (1 means perfect).

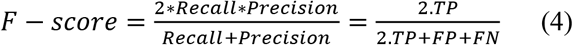
iii. ***Kappa***. Kappa is a coefficient developed to measure observed agreement normalized to the agreement by chance.

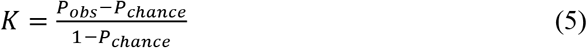
iv. ***Sensitivity/Recall:*** Sensitivity is defined as the proportion of positive instances that are correctly classified as positive, with respect to all positive set.

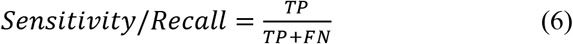
v. ***Specificity:*** Specificity is the proportion negative instances that are correctly identified as negative out of the total observed negative instances

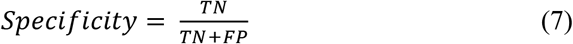
vi. ***Precision:*** Precision is the true positive divided by the sum of all positive prediction

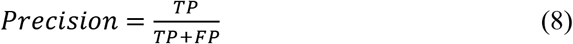
vii. ***ROC-AUC:*** An ROC stands for Receiver Operating Characteristics is graphical plot illustrate binary classifier performance for various thresholds. ROC is created by plotting the true positive rate (TPR) against the false positive rate (FPR) at various threshold settings [23]. The area under the ROC curve (AUC) summarized the ROC performance to single value, it ranged between 0 and 1. A higher AUC value represents the superiority of a classifier and vice versa [14]

## 3. Results

The mean age of the patients was 53.1 years old (the median was 54) and majority (61.2%) of them were male Overall, 38.4% (163) of patients were died and 61.6% (262) discharged alive.

As shown in Table 2 above and visualized in Fig. 1 below, 3.3% (14) of the patients were less than 20 years of age and all of them survived the disease, 28.5% (121) aged between 20 to 39 and 27.8% (118) of them discharged alive, 27.5% (117) were between 40 to 59 years old of which 76.9% (90) of them discharged from hospital, 28.5% (121) of the patients were within 60 to 79 years of age and only 31,4% (38) of them survived, lastly 12.2% (52) of the patients aged 80 years and above of which 96.2% (50) of them died for COVID-19. The result for test of significant difference between age group and survival outcome, indicates p-value =0.000 (<0.05). Hence, the null hypothesis is rejected and conclude that the two variables (age and patients’ survival outcome) relate to each other (dependent). As for distribution of the sex of the patients, 33.8% of the patients were female and 63.5% of them were discharged while 60.4% of the male survived. The chi-square test result for Sex and patient’s outcome has P-value = 0.5690 (> 0.05) which indicates the two variables are in fact independent.

**Table 2:** Distribution of demographic and clinical characteristics of COVID-19 patients with the survival outcome.

| Demographics and clinical Variables |  | Total | Died (n=163) | Discharged (n=262) | P-value |
| --- | --- | --- | --- | --- | --- |
| Age group | 0-19 years | 14 (3.3%) | 0 | 14 (3.3) | 0.000 |
|  | 20-39 years | 121 (28.5) | 3 (0.7) | 118 (27.8) |  |
|  | 40-59 years | 117 (27.5) | 27 (6.4) | 90 (21.2) |  |
|  | 60-79 | 121 (28.5) | 83 (19.6) | 28 (8.9) |  |
|  | 80+ years | 52 (12.2) | 50 (11.8) | 2 (0.5) |  |
| **Sex** | Female | 165 (38.5) | 60 (36.4) | 105 (63.6) | 0.502 |
|  | Male | 260 (61.2) | 103 (39.6) | 157 (60.4) |  |
| **Symptoms** | arrhythmia | 1 (1.2) | 1 (100) | 0 | 0.000 |
|  | acute respiratory distress | 50 (11.8) | 50 (100) | 0 |  |
|  | arrhythmia | 4 (0.9) | 4 (100) | 0 |  |
|  | cough & fever | 28 (6.6) | 6 (21.4) | 22 (78.6) |  |
|  | diarrhea | 2 (0.5) | 0 | 2 (100) |  |
|  | difficulty breathing | 2 (0.5) | 2 (100) | 0 |  |
|  | discomfort | 2 (0.5) | 0 | 2 (100) |  |
|  | dry cough | 1 (0.2) | 1 (100) | 0 |  |
|  | fever & fatigue | 27 (6.4) | 9 (33.3) | 18 (66.7) |  |
|  | myelofibrosis | 1 (0.2) | 1 (100) | 0 |  |
|  | pneumonia | 25 (5.9) | 25 (100) | 0 |  |
|  | respiratory symptoms | 2 (0.5) | 0 | 2 (100) |  |
|  | sore throat | 2 (0.5) | 1 (50) | 1 (50) |  |
|  | NONE | 278 (63.4) | 63 (22.7) | 215 (77.3) |  |
| **Travel history** | NO | 333 (78.4) | 157 (47.1) | 176 (52.9) | 0.000 |
|  | YES | 92 (21.6) | 6 (6.5) | 86 (96.5) |  |
| **Comorbidity (binary)** | NO | 317 (74.5) | 75 (23.7) | 242 (76.3) | 0.000 |
|  | YES | 108 (25.4) | 88 (81.5) | 20 (18.5) |  |
| **Comorbidity** | Asthma | 5 (1.2) | 2 (40) | 3 (60) | 0.000 |
|  | cardiac disease | 1 (0.2) | 1 (100) | 0 |  |
|  | chronic kidney disease | 7 (1.6) | 7 (100) | 0 |  |
|  | COPD | 2 (0.5) | 0 | 2 (100) |  |
|  | diabetes | 22 (5.2) | 22 (100) | 0 |  |
|  | hepatitis B | 2 (0.5) | 2 (100) | 0 |  |
|  | hypertension | 61 (14.4) | 49 (80.3) | 12 (19.7) |  |
|  | prostate cancer | 2 (0.5) | 2 (100) | 0 |  |
|  | prostate hypertrophy | 2 (0.5) | 0 | 2 (100) |  |
|  | pulmonary disease | 2 (0.5) | 2 (100) | 0 |  |
|  | NONE | 319 (75.1) | 76 (23.8) | 243 (76.2) |  |
| **LOS** | Above | 186 (43.8) | 6 (3.2) | 180 (96.8) | 0.000 |
|  | Below | 239 (56.2) | 157 (65.7) | 82 (34.3) |  |

**Fig 1:**
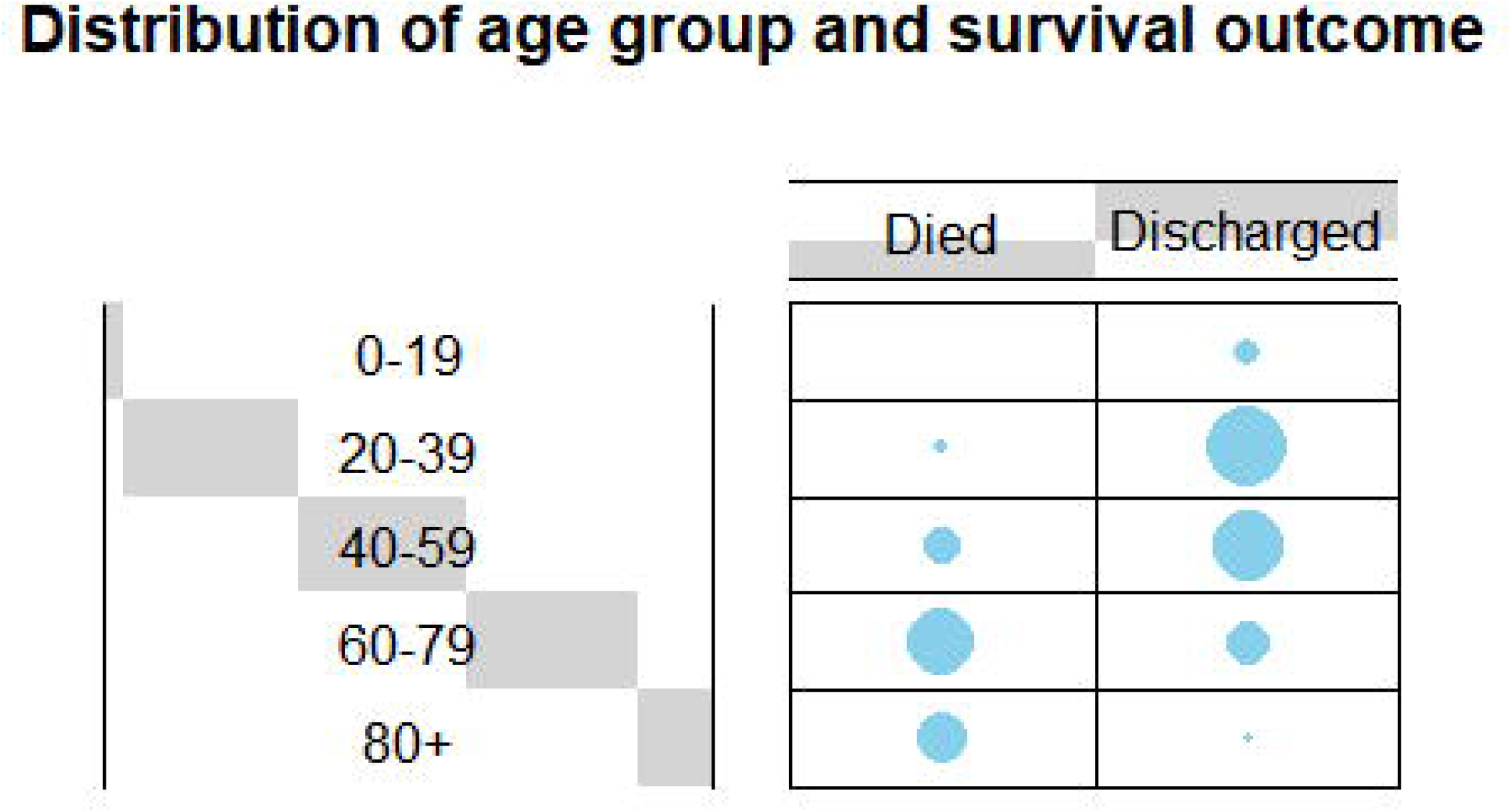
Distribution of age group and survival outcome

Symptoms of the patients on admission as shown in Table 2, 63.4% (278/425) of the patient are asymptotic while 147 have shown at least one symptoms and the most commonly experienced symptoms were acute respiratory distress 34%(50/147) followed by cough & fever 19.1% (28/147), and fever & fatigue 18.4%(27/147). On the relationship between onset symptoms and survival status, 77.3% (215) of patients reported no symptoms on admission survived the disease. All patients with acute respiratory symptoms died and more than 70% of patients with onset fever were discharged. A similar result was illustrated in Fig. 2. The associativity test between symptoms and survival outcome result has P-value = 0.000 (P<0.05) hence, the two variables are statistically significantly related.

**Fig 2:**
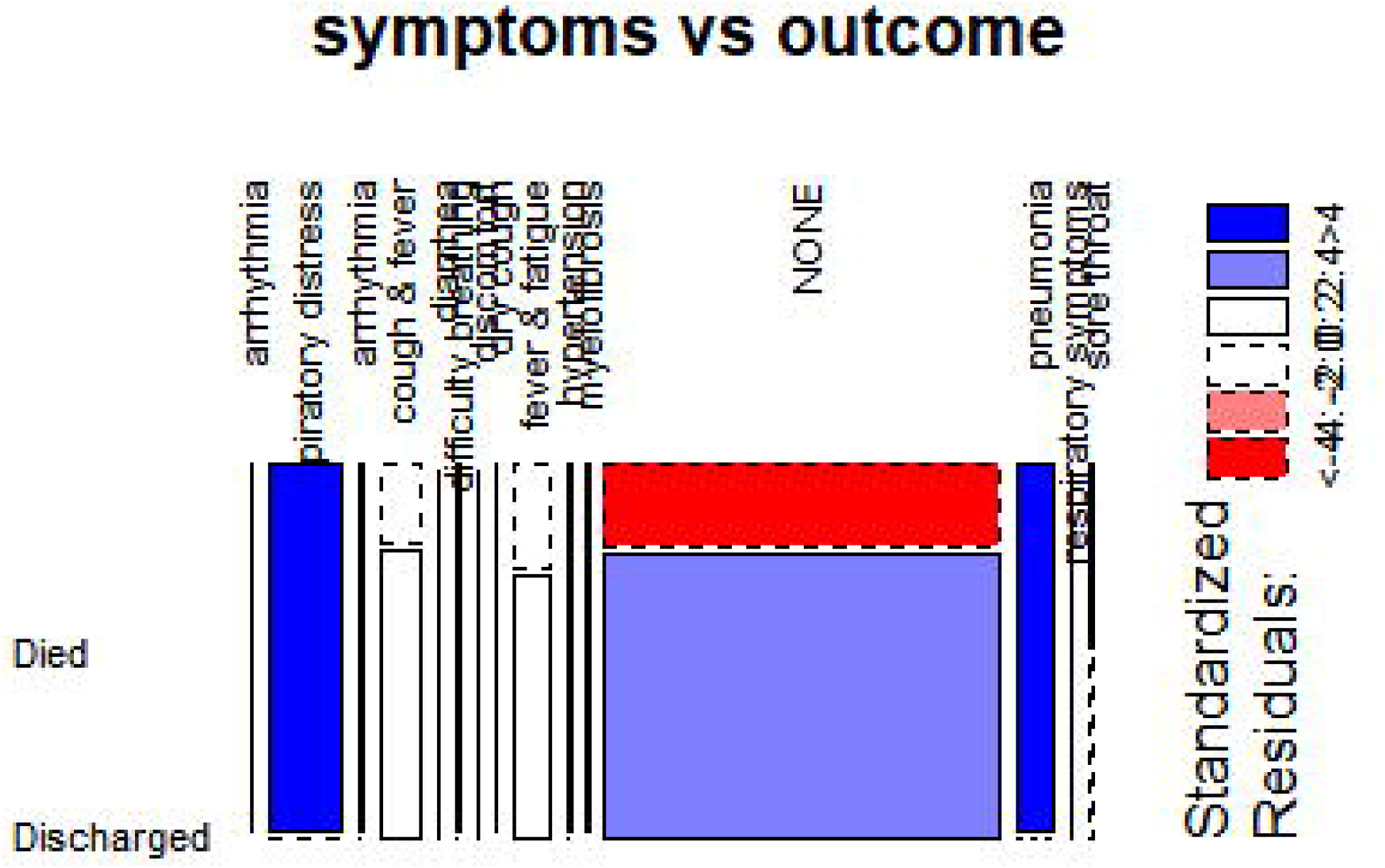
Distribution of symptoms and survival outcome

The distribution of travel history and survival status of COVID-! 19 patients are as follows. 78.4% of the patient has no travel history and 52,9% of them discharged. The mortality rate of a patient with travel history is 6.5%. the chi-square result showed the emergence of a significant association between travel history and survival outcome with p-value <0.05.

The comorbidity was in this case treated as binary (YES versus NO) before subsequently, grouped based on specific comorbidity. [3] point out that both the dummy variable and specific comorbidity could be simultaneously used in predicting COVID-19 patients’ survival outcome. Results from cross-tabulation indicates 25.4% of the patients under study have a history of underline medical problems of whom only 18.5% of them survived and 81.5% were died which indicates very high mortality rate of patients with comorbidity. The associativity test of chi-square reveals dependency between Chronic diseases and survival outcome (P-value=0.000) and Fig.3 explains further the nature of associativity, as we can see having comorbidity (YES) is inversely associated with “Discharged” and positively correlated with “Death”. This means those with comorbidity diseases are more likely to die with COVID-19 while those without it have a higher chance of recovering from the disease.

**Fig. 3.**
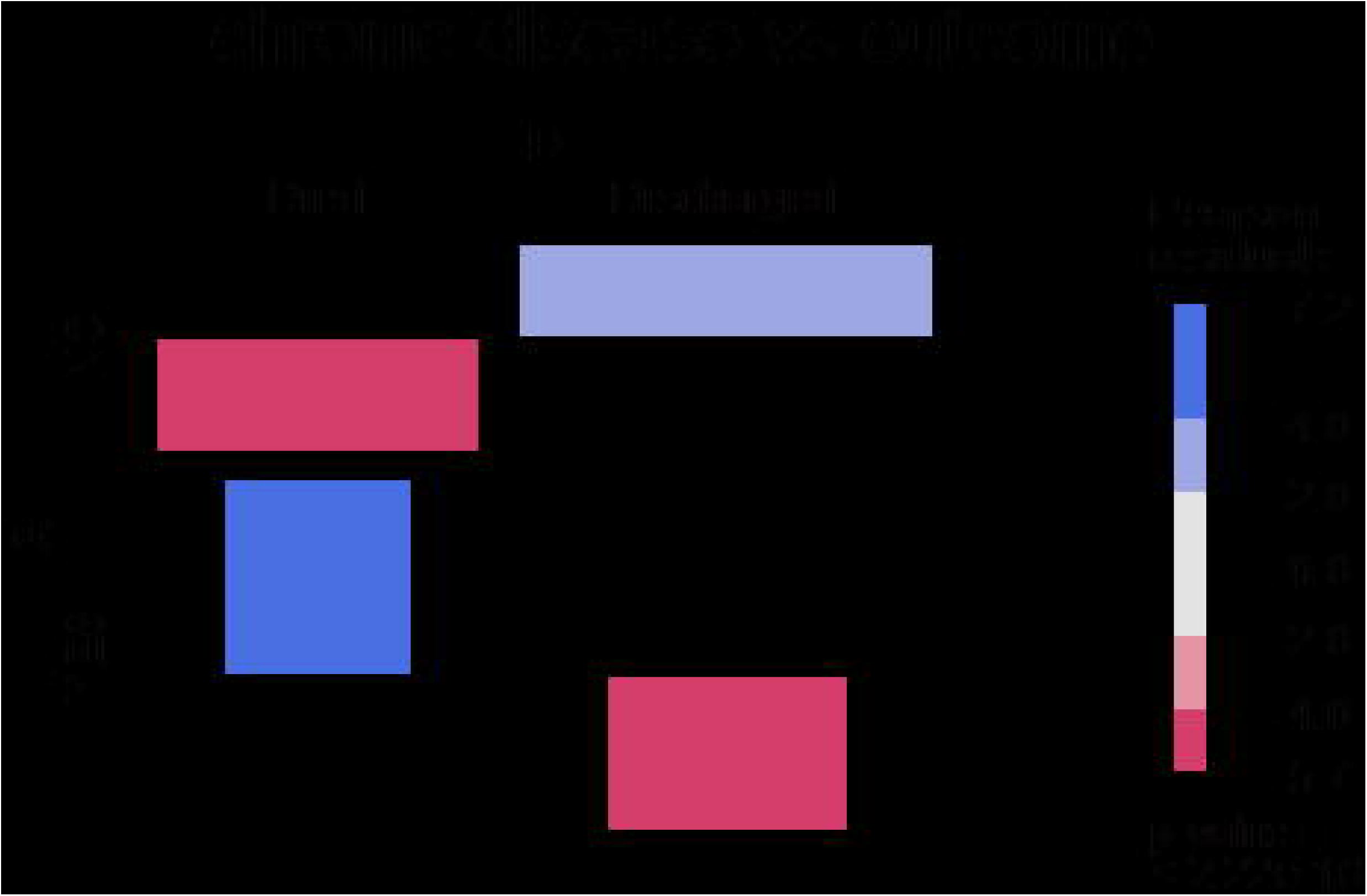
Distribution of comorbidity (binary) and outcome

Table 2 also expressed the distribution of specific comorbidities recorded on admission. 106 (24.1%) patients had at least one underlying comorbidity, the most frequent of which were chronic diseases, such as hypertension (57.5%) and diabetes (20.6%), only two COPD patients were identified. All patients with diabetes and kidney disease died on admission while more than 80% of the patients with hypertension died. Out of 425 patients, 75.1% (319) had no documented comorbidity and 76.2% (243) of them were discharged alive. The Chi-square test results describe the specific comorbidities and survival outcome to have a statistical significant relationship

Hospital Length of Stay (LOS) was converted from numerical to dummy variable using the average length of stay (ALOS) which was calculated to be 9 days, all the patients stayed above average were tagged “A” and those stayed below average tagged “B”. Table 2 shows the distribution of the stratified LOS and survival outcome, among 186 patients stayed above ALOS 180 (96.8%) were discharged. Death was higher in patients stayed below ALOS, 157 (65.7) out of 239 of them died. The results can be visualized from Fig. 4 which indicated the longer patient stayed the more likely he is to survive the disease.

**Fig. 4.**
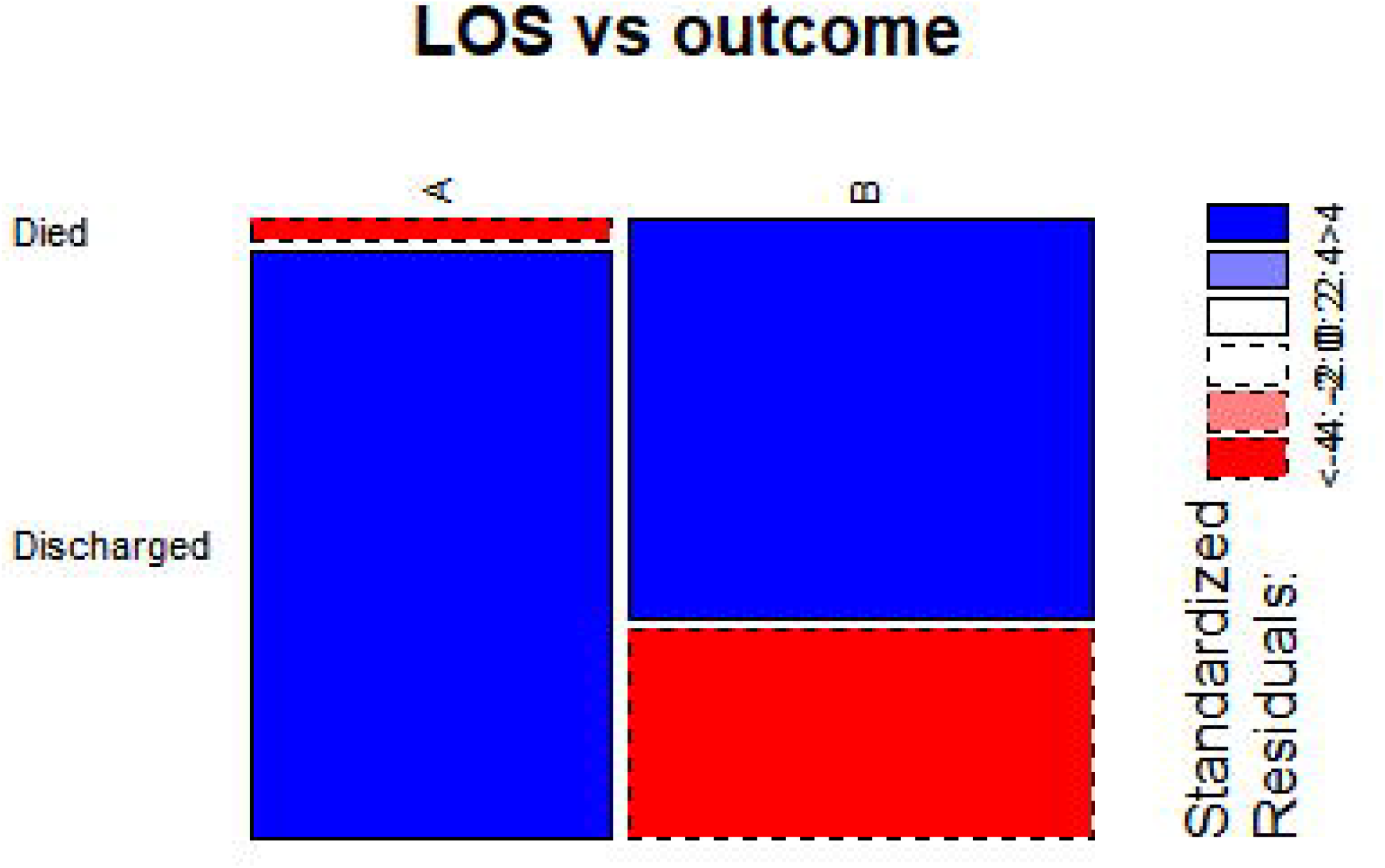
Distribution of length of stay and survival outcome

### 3.1 Linear Discriminant Analysis

Confusion matrix was utilized to visualize the performance of the machine learning algorithms as on test data for which the true values are known after building the models using the training set. Confusion matrix outcome was used to calculate model performance evaluation metrics such as accuracy, precision and soon.

Table 3 represents the confusion matrix for the LDA model, as it can be seen that the model was able to predict 30 dead patients correctly and misclassified 2 as discharged, also correctly classified 50 discharged patients and misclassified 2. The model has an overall classification accuracy of 0.95 (95%).

**Table 3:** Confusion matrix for LDA.

| Predicted classes | Observed classes |  |  |
| --- | --- | --- | --- |
|  | Died | Discharged | Row Total |
| Died | 30 | 2 | **32** |
| Discharged | 2 | 50 | **52** |
| Column Total | **32** | **52** | **84** |

### 3.2 Random Forest

Table 4. showed model performance for the random forest classifier using the confusion matrix. The model appears to perform perfectly by correctly classifying all 32 dead patients and classified all 52 discharged patients correctly without any misclassification, leading to the overall classification accuracy of 100% and therefore the error rate is 0%. The importance of each predictor in determining the survival outcome of CIVID-19 patients with a random forest model was also explored and the result was presented in Fig. 5, LOS is the most important variable followed by age then symptoms using both Mean Decrease Accuracy and Mean Decrease Gini. Sex is a less important variable.

**Table 4:**
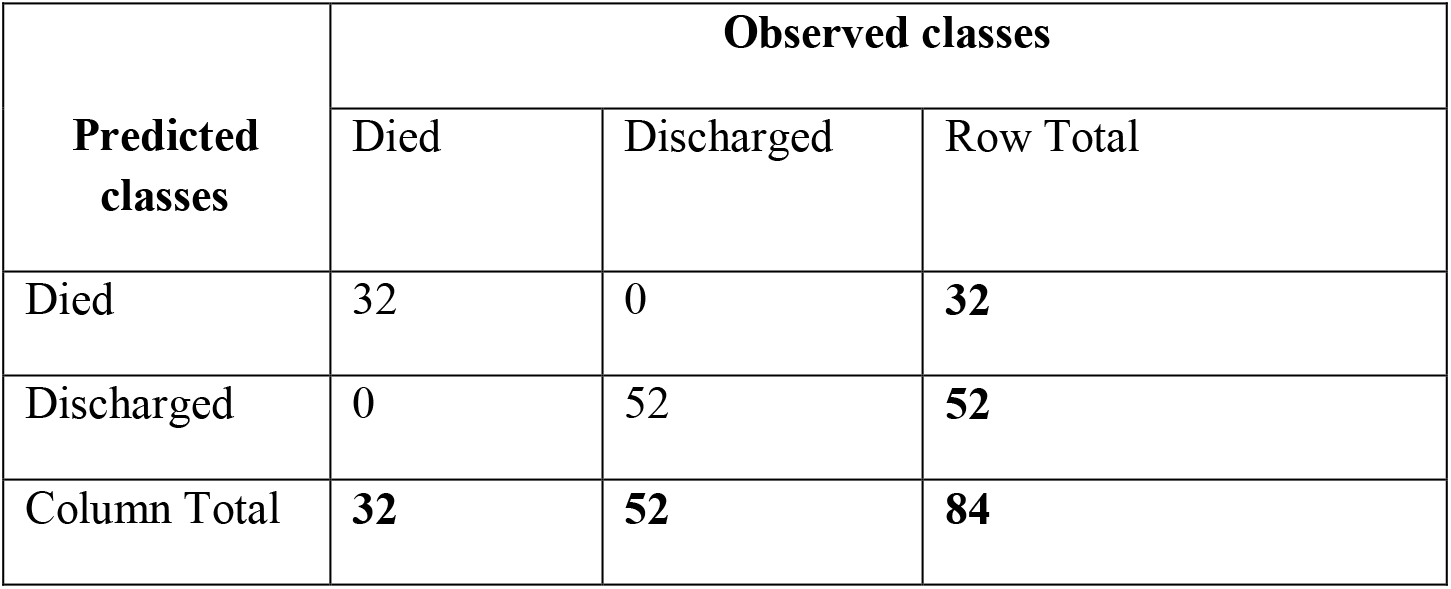
Confusion matrix for RF.

| Predicted classes | Observed classes |  |  |
| --- | --- | --- | --- |
|  | Died | Discharged | Row Total |
| Died | 32 | 0 | **32** |
| Discharged | 0 | 52 | **52** |
| Column Total | **32** | **52** | **84** |

**Figure 5:**
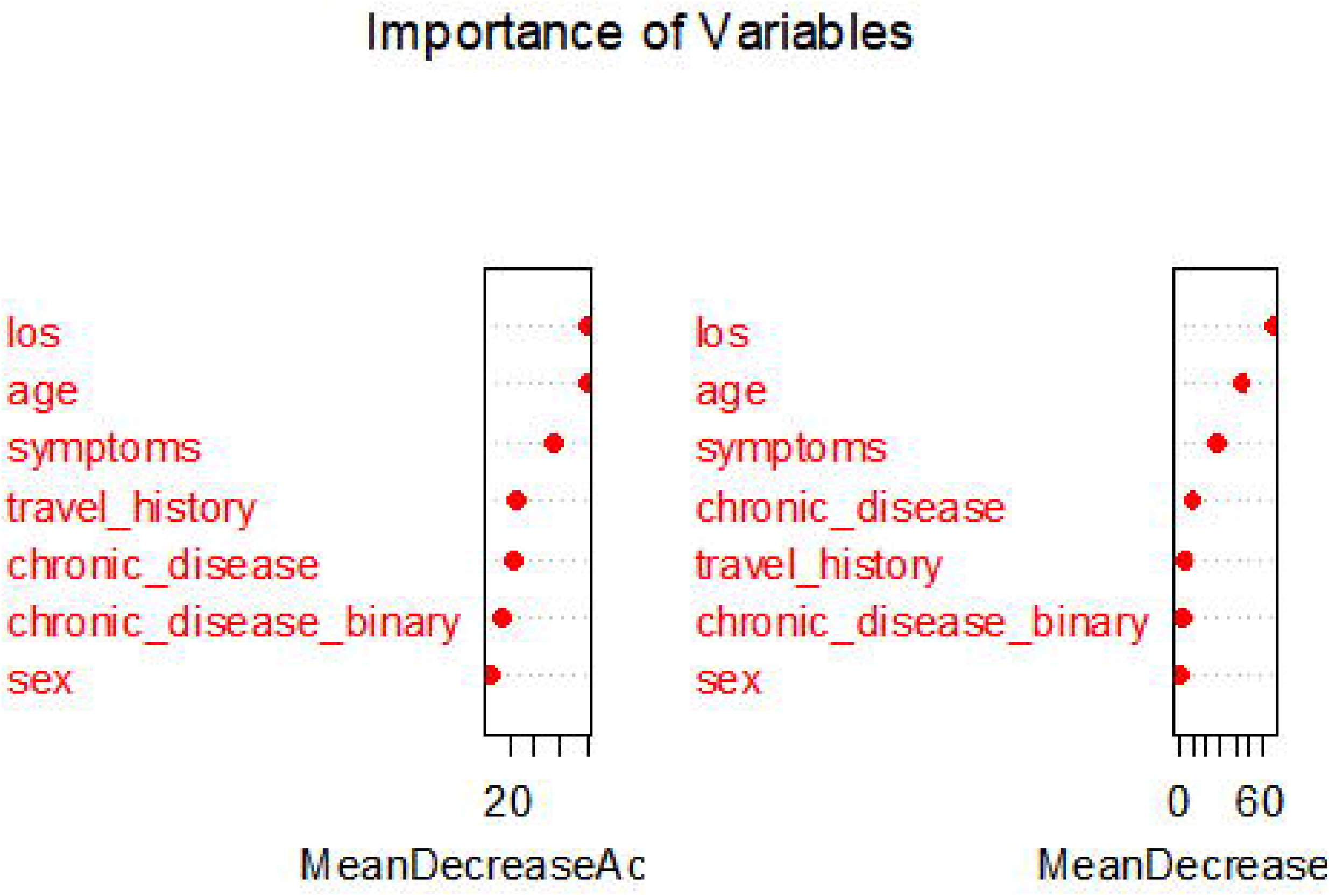
plot for variable importance

### 3.3 Support Vector Machine

Let’s have a look at the results of the performance of the SVM algorithm on the test dataset in Table 5, in which 30 out of 32 died patients are correctly classified and 2 were misclassified while 49 out of 52 discharged patients were correctly classified. The overall classification accuracy was 94% and the apparent error rate of 6%.

**Table 5:**
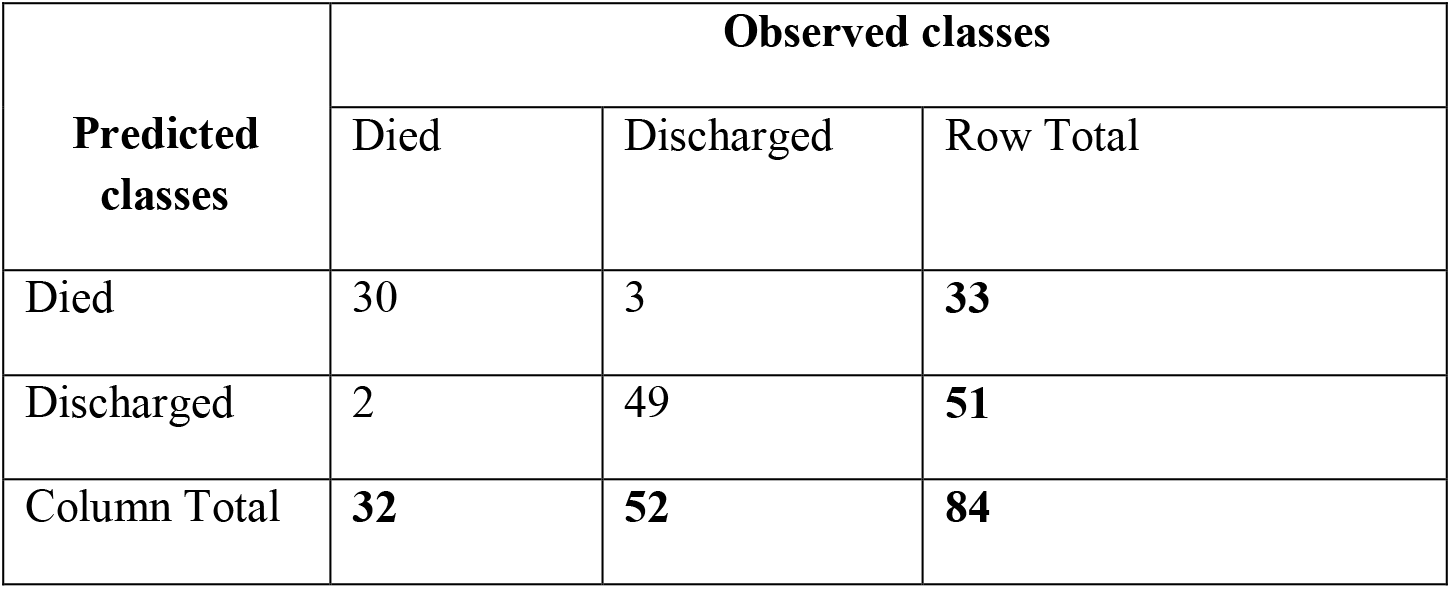
Confusion matrix for SVM

### 3.4 Performance comparison of the models

We compared the classification accuracy of the three different supervised machine learning algorithms namely, LDA, RF, and SVM. All of the algorithms performed absolutely well by achieving greater than 90% accuracy. The recall and precision rates for each of the algorithms also showed a similar result to that seen with overall accuracy. Table 6 shows the predictive performances of the classifiers for the test data set. It is observed that RF have better performance on all evaluation metrics: accuracy (100%), precision, recall, specificity, F-score, Kappa and AUC, followed by LDA which also performed very well, its prediction accuracy has been calculated as 95.2% of accuracy, 93.8% recall, 96.2% specificity, and 89.9% Kappa index. Fig. 7 illustrates the ROC graph for the three classifiers (i.e., LDA, RF, and SVM). Figure 6 shows three ROC curves for all the three developed models based on the given outcome. RF has outperformed the other classifiers with an area under the curve(AUC) 0.994. Similarly, LDA and SVM perform excellently with the area under the curve(AUC) 0.991 and 0.970 respectively.

**Table 6.**
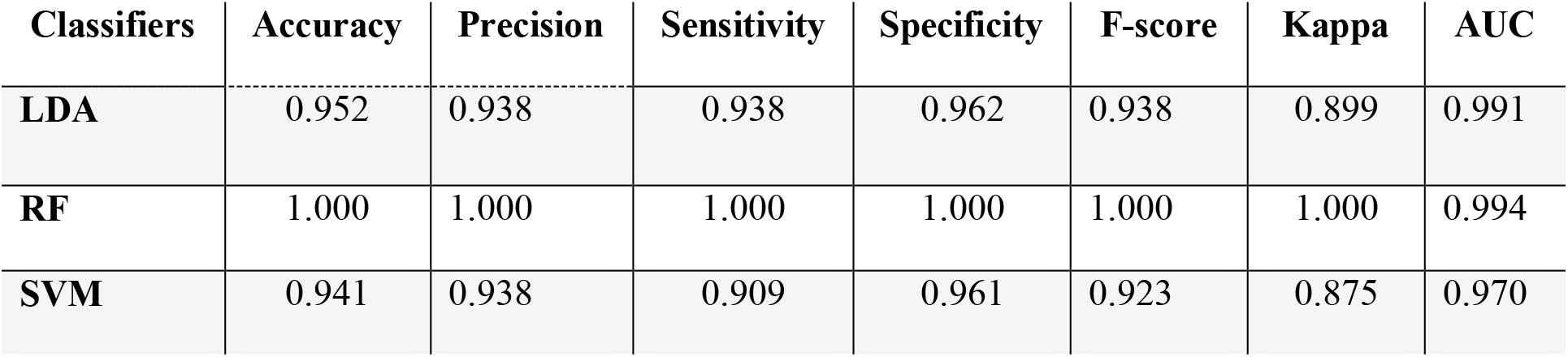
Summary of model performance for the three classifiers

| Classifiers | Accuracy | Precision | Sensitivity | Specificity | F-score | Kappa | AUC |
| --- | --- | --- | --- | --- | --- | --- | --- |
| **LDA** | 0.952 | 0.938 | 0.938 | 0.962 | 0.938 | 0.899 | 0.991 |
| **RF** | 1.000 | 1.000 | 1.000 | 1.000 | 1.000 | 1.000 | 0.994 |
| **SVM** | 0.941 | 0.938 | 0.909 | 0.961 | 0.923 | 0.875 | 0.970 |

Figure 6. ROC curve for LDA, RF and SVM on the task of classifying survival outcome of COVID-19 patients

## Discussion

This study explored factors influencing survival outcomes of COVID-19 patients. The mean age of all patients was 53.1 years (the median was 54) which is close to that of data reported by [4] [24] 56.0 years and [10] 57 years, older than 48.9 years reported by [3] and, but younger than that reported by [11] 73 years and [6] 70.6 years. Chi-square test was used to test the associativity between the six predictors and survival outcome. Table 2 shows the results considering a significant level of 0.05. Age was discovered to be highly associated with the survival outcome, it was found that those with older age had a higher likelihood to die of COVID-19 than those with younger age. the results also indicate independence between sex and survival outcome. Age has previously reported by many researchers including [24] [25] [11] to be associated with the death of patients with COVID-19. [25] reported sex not have significance in determining patient’s outcome. Results of this analysis as displayed in Table 2 shows that 63.4% (278) of patients reported no symptoms on admission. The most frequent symptoms were acute respiratory distress, cough, fever and fatigue. [4] [5] [3] highlighted fever, fatigue and dry cough as the most common COVID-19 symptoms. It is further observed that patient with onset symptoms are more likely to die of the disease. Also patients with acute respiratory distress and pneumonia are most vulnerable to severe condition that could lead to death. The finding on patients with morbidity is that out of the 425 patients we studied 108 (25.4%) have reported at least one morbidity on admission and only less than 20% of them discharged alive. Hypertension (57.5%) and diabetes (20.6%) were the most common underlying diseases and recorded higher mortality rates. These supported the [3] [5] [4] [6] findings. Hospital Length of Stay (LOS) was found to influence the patient’s survival outcome, Table 2 shows that 96.8% of patients stayed longer than the average length of stay survived. Hence, longer LOS increases the chances of patients been discharged alive.

The study developed a machine learning techniques for prediction of COVID-19 patient’s survival outcome after a hospital stay, using the patients’ demographic and clinical characteristics believed to contributed to the outcomes. Three methods LDA, RF, and SVM were applied on the COVID-19 data set, our results were presented in Tables 3-5 using a confusion matrix which determines classifiers performance based on classified instances. According to Table 3 LDA correctly classified 80 instances out of 84, yielding accuracy of 95.4%. The performance of RF was presented in Table 4, this algorithm correctly classified all the instances with zero misclassification. Table 5 shows the performance of SVM algorithms which classified 79 instances correctly out of a total number of 84 instances having an accuracy of 90.9%. From these we can conclude that based on accuracy RF classifier presented a higher performance in comparison with the others.

The general performances of the models were examined based on seven performance metrics. consider Table 6, it clearly shown that RF has highest accuracy (100%), recall (100%), precision (1.00) and specificity (1.00) when comparing with LDA who has Accuracy (0.952), recall (0.938), precision (0.938) and specificity (0.962) and SVM has Accuracy (0.941), recall (0.909), precision (0.983) and specificity (0.961), we can clearly see that RF is the best model while LDA is the better then SVM. Based on the Kappa Statistic which is used to assess the accuracy of any particular measuring cases, RF has the highest value of 1.00 followed by LDA (0.899) then SVM (0.875). ROC curve represents the combination of sensitivity and specificity. Theoretically, the area under the ROC (AUC) can assume values between 0 and 1, where an ideal classifier will take the value of 1. However, the practical lower bound for random classification is 0.5 which means the classifier with no discriminative capability. Whereas classifiers with an AUC significantly higher than 0.5 have at least some ability to discriminate. the experimental results of the study were displayed in Figure 4 for LDA, RF, and SVM. All the algorithms did well but the AUC for RF is 0.994, showing the reliability of discriminative capability among all the methods. We can conclude that both LDA, RF and SVM perform outstandingly. However, the RF algorithm is considered the best supervised machine learning algorithms of this study.

## Conclusions

This paper presents a comparative studies of three machine learning algorithms to predict of the survival outcomes of COVID-19 patients. The algorithms were evaluated on seven different performance metrics (accuracy, sensitivity, specificity, and precision, recall, F-measure, ROC and AUC). The results shown solid prediction capabilities of Machine Learning Techniques in prediction of COVID-19 patients survival status, all the algorithms performed very well but the RF classifier are considered as best model. The study also demonstrates that, patients with underline comorbidities and aging patient are more likely to develop severe situation, and have little chance of surviving COVID-19. In terms of duration of hospital stay the longer patient stays the higher chance of his surviving. Early detection COVID-19 patients is essential for identification of vulnerable patients who may need special care to survive the disease, optimal usage of resources as well as estimation of number of beds required in intensive care units.

## Data Availability

Data used was extracted from https://drive.google.com/file/d/1bYcMAd-lOiSlOVGl5qxG1Sq1auyzmWHZ/view, obtained from various government agencies and compiled by Dr. Bharatendra Rai, Director, Master of Science Technology Management Program and Chair, Dept. of Decision & Info Sciences at UMass Dartmouth.

